# Estimating the impact of reopening schools on the reproduction number of SARS-CoV-2 in England, using weekly contact survey data

**DOI:** 10.1101/2021.03.06.21252964

**Authors:** James D Munday, Christopher I Jarvis, Amy Gimma, Kerry LM Wong, Kevin van Zandvoort, CMMID COVID-19 Working Group, Sebastian Funk, W. John Edmunds

## Abstract

**Background:** Schools have been closed in England since the 4th of January 2021 as part of the national restrictions to curb transmission of SARS-CoV-2. The UK Government plans to reopen schools on the 8th of March. Although there is evidence of lower individual-level transmission risk amongst children compared to adults, the combined effects of this with increased contact rates in school settings are not clear.

**Methods:** We measured social contacts when schools were both open or closed, amongst other restrictions. We combined these data with estimates of the susceptibility and infectiousness of children compared with adults to estimate the impact of reopening schools on the reproduction number.

**Results:** Our results suggest that reopening all schools could increase R from an assumed baseline of 0.8 to between 1.0 and 1.5, or to between 0.9 and 1.2 reopening primary or secondary schools alone.

**Conclusion:** Our results suggest that reopening schools is likely to halt the fall in cases observed in recent months and risks returning to rising infections, but these estimates rely heavily on the current estimates or reproduction number and the current validity of the susceptibility and infectiousness profiles we use.

## Introduction

School closures have been implemented in many countries as part of a broader response to the COVID-19 pandemic [1]. It is well established that children are at low risk of hospitalisation and death as a direct result of infection [2, 3]. Despite this lower risk, there is concern that allowing transmission amongst younger age-groups increases risk of infection in adults, who are at substantially higher risk. The role of schools in transmission is therefore an important question. On the 4th of January 2021, a third national lockdown was announced in England to curb transmission of SARS-CoV-2 [4]. This included the closure of schools, a measure the UK government plans to reverse on the 8th of March.

The direct and indirect impact of school closures and eventual reopening is still unclear. There is mixed evidence around the role of schools in community transmission. Existing studies of transmission within schools have wide ranging results [5–7]. Other work demonstrates an increased prevalence amongst school-aged-children when schools return [8, 9] and a higher risk of infections entering households through children than adults. However, the evidence that schools drive transmission in the community remains scarce [10, 11]. A particular challenge for many analyses is bias resulting from the age-dependence in case ascertainment due to varying rates of asymptomatic infection [12]. This challenge is then further complicated by changes in epidemiology due to the emergence of new variants [13].

The potential change in transmission of SARS-CoV-2 upon reopening schools predominantly depends on a combination of two factors. Firstly, the age-specific risk of transmission upon contact. Secondly, the likely increased rate of contact between members of the population due to school reopening. Multiple studies aimed at understanding the relative transmission risk associated with children indicate lower susceptibility [14–16] and some indicate lower infectiousness [14]. However, evidence of lower transmission risk amongst children alone is insufficient to quantify the impact of reopening schools. There is a need to combine the estimates of reduced susceptibility and infectiousness with age specific contact patterns in this age-group social contacts amongst school-aged-children.

There is abundant evidence that children’s contacts increase when schools are open, presenting opportunities for increased infectious disease transmission which is well documented in other pathogens such as influenza [17]. Nonetheless, it is important to capture how these contacts vary under the specific conditions presented during the current pandemic response, where social distancing and other mitigations are in effect within schools.

CoMix is a large-scale comprehensive social contact survey which has collected data on social contacts in the UK on a weekly basis since the 24th of March 2020 [18]. In this paper, we estimate the impact of opening schools on the reproduction number in England, by combining social-contact data collected during periods where schools were open and closed [18] with estimates of age-stratified susceptibility and infectiousness [14–16].

## Methods

### CoMix Data

CoMix is a longitudinal behavioural survey, launched on the 24^th^ of March 2020. The sample is broadly representative of the UK adult population with data collected from approximately 2000 individuals per week. Participants are invited to respond to the survey once every two weeks. We collected weekly data by running two alternating panels. Parents complete the survey on behalf of children (17 years old or younger). Participants record direct, face-to-face contacts made on the previous day, specifying certain characteristics for each contact including the age and sex of the contact, whether contact was physical (skin-to-skin contact), and where the contact occurred (e.g. at home, work, while undertaking leisure activities, etc). Further details have been published elsewhere [18]. The contact survey is based on an approach developed for the POLYMOD contact survey [19]. We provide a brief descriptive analysis of the contacts recorded during the November and January lockdown periods by age group and geographical region.

### Constructing contact matrices and estimating reproduction number

We constructed age-stratified contact matrices for nine age-groups (0-4, 5-11, 12-17, 18-29, 30-39, 40-49, 50-59, 60-69, and 70+). Participants did not report exact ages of contacts, we therefore sampled from the reported age-group with a weighting consistent with contacts reported in the POLYMOD survey. We fitted a truncated negative binomial model to calculate the mean contacts between each participant and contact age-groups. To ensure reciprocity in contacts, we multiplied the matrix by population size vector for England, using United Nations World Population Prospects data [20], before taking the cross-diagonal mean and then dividing by the same population vector again.

### Profiles of Age-dependent transmission risk

We consider five age-dependent susceptibility and infectiousness profiles (Table 1):

**Table 1.** Susceptibility and infectiousness profiles taken from Davies et.al.[14], ONS reports and Viner et al[16]

| Study | Age groups | Susceptibility | Infectiousness | Clinical Fraction |
| --- | --- | --- | --- | --- |
| Davies et al<sup>1</sup> | 0-4 | 0.4 (0.25, 0.57) | 0.61 | 0.29 (0.18, 0.44) |
|  | 5-10 | 0.4 (0.25, 0.57) | 0.61 | 0.29 (0.18, 0.44) |
|  | 11-17 | 0.4 (0.27, 0.53) | 0.61 | 0.21 (0.12, 0.31) |
|  | 18-29 | 0.79 (0.59, 0.96) | 0.64 | 0.27 (0.18, 0.38) |
|  | 30-39 | 0.86 (0.69, 0.98) | 0.67 | 0.33 (0.24, 0.43) |
|  | 40-49 | 0.80 (0.61, 0.96) | 0.70 | 0.40 (0.28, 0.52) |
|  | 50-59 | 0.82 (0.63, 0.97) | 0.75 | 0.49 (0.37, 0.60) |
|  | 60-69 | 0.88 (0.70, 0.99) | 0.82 | 0.63 (0.49, 0.76) |
|  | 70+ | 0.74 (0.56, 0.90) | 0.85 | 0.69 (0.57, 0.82) |
|  |  | Susceptibility | Infectiousness |  |
| ONS<sup>2</sup> | 0-4 | 0.5 (0.35, 0.75) | 1.0 (0.7, 1.5) |  |
|  | 5-10 | 0.5 (0.35, 0.75) | 1.0 (0.7, 1.5) |  |
|  | 11-17 | 0.5 (0.35, 0.75) | 1.0 (0.7, 1.5) |  |
|  | 18-29 | 1.0 | 1.0 |  |
|  | 30-39 | 1.0 | 1.0 |  |
|  | 40-49 | 1.0 | 1.0 |  |
|  | 50-59 | 1.0 | 1.0 |  |
|  | 60-69 | 1.0 | 1.0 |  |
|  | 70+ | 1.0 | 1.0 |  |
|  |  | Susceptibility | Infectiousness |  |
| Viner et al<sup>3</sup> | 0-4 | 0.64 (0.51, 0.81) | 1.0 (assumed) |  |
|  | 5-10 | 0.64 (0.51, 0.81) | 1.0 (assumed) |  |
|  | 11-17 | 0.64 (0.51, 0.81) | 1.0 (assumed) |  |
|  | 18-29 | 1.0 | 1.0 |  |
|  | 30-39 | 1.0 | 1.0 |  |
|  | 40-49 | 1.0 | 1.0 |  |
|  | 50-59 | 1.0 | 1.0 |  |
|  | 60-69 | 1.0 | 1.0 |  |
|  | 70+ | 1.0 | 1.0 |  |
|  |  | Susceptibility | Infectiousness |  |
| CoMix fit | 0-4 | 0.31 (0.30, 0.31) | 1.0 |  |
|  | 5-10 | 0.31 (0.30, 0.31) | 1.0 |  |
|  | 11-17 | 0.31 (0.30, 0.31) | 1.0 |  |
|  | 18-29 | 1.0 | 1.0 |  |
|  | 30-39 | 1.0 | 1.0 |  |
|  | 40-49 | 1.0 | 1.0 |  |
|  | 50-59 | 1.0 | 1.0 |  |
|  | 60-69 | 1.0 | 1.0 |  |
|  | 70+ | 1.0 | 1.0 |  |

<sup>1</sup> 95% Credible Intervals
<sup>2</sup> Approximate results inferred from plot in[15] unknown quantification of uncertainty
<sup>3</sup> 95% Confidence Interval

The first profile (i) assumed equal susceptibility and infectiousness in all age groups. This is unlikely to reflect reality but provides an upper limit as a reference point to compare the other profiles.

For the second profile (ii) we used results from a mathematical modelling study by Davies et. al [14]. which estimated relative susceptibility and clinical fraction in 9 age groups. The work also reports estimates of 50% infectiousness of sub-clinical cases and reports clinical fraction by age. We used this to calculate infectiousness per age group further detailed in Table 1.

The third profile (iii), was based on analyses of household transmission patterns from the Office for National Statistics (ONS) Community Infection Study [15]; 50% susceptibility in children relative to adults but equal infectiousness.

For the fourth profile (iv), we performed a meta-analysis of prevalence studies included in a systematic review by Viner et al [16]. We used a random effects model based on the data from Figure 4 of their paper. This resulted in 64% (51% - 81%, 95% confidence interval [CI]) susceptibility in children relative to adults, we assumed equal infectiousness between children and adults [16];

For the fifth profile (v), we used an independent estimate of relative susceptibility in children (31%, see results section), quantified by comparing reproduction numbers estimated from CoMix data and using a well-established time-series method developed by Abbott et. al [21], which uses a time-series of cases to determine the instantaneous reproduction number under an assumed generation interval and infection to reporting delay distribution.

### Inferring age dependent transmission risk using CoMix data

We established independent estimates of susceptibility and infectiousness in children relative to adults. We did this by comparing estimates of *R* using CoMix contact data with estimates of the time-varying reproduction number in England calculated using case data [21]. To capture the change in contact rates as schools returned in September 2020

We calculated a reproduction number resulting from two-weekly rolling contact matrices **C**_*t*_and assumed relative susceptibility and infectiousness vectors **s** and **i** to be:

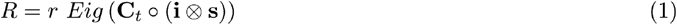

We simplified **s** and **i** such that adult age-groups (18+) were 1.0 and child age groups were equal, *s* and *i*. We inferred *s* and *r*, keeping *i* at 1.0, by fitting our estimates using maximum likelihood estimation to those calculated using the EpiNow2 package [21]. We assumed gamma distributed uncertainty in the time-varying estimates which we parameterised using the mean μ_*rt*_ and standard deviation σ_*rt*_ of these estimates over each survey period used to calculate CoMix derived eigenvalues.

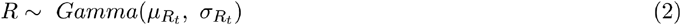

To show the likelihood surface of relative susceptibility and infectiousness, we calculated the likelihood of a range of combinations of *i* and *s* while fitting *r*.

We fitted over 2 periods of time. Firstly, between 27th July and 10th October to most clearly capture the impact of schools returning in the summer whilst minimising issues related to gradual acquisition of natural immunity. Second, We fitted over a longer period of time incorporating data from 10th June.

We omitted data at the end of August in both fits due to a short spike in reproduction number estimates, which we believe resulted from large numbers of imported cases from recreational travel. We further omitted two weeks in July when contacts were not recorded for children. We assessed sensitivity to the fitted period, by using a range of fitting options (Figure S4).

### Evaluating the impact of reopening schools on Reproduction Number

We created contact matrices using CoMix data collected during the second lockdown, (5th November to 2nd December 2020) to represent contacts during a lockdown with schools open. We used data from 5th to 18th of January 2021 for contacts during a lockdown with schools closed (Supplementary Figures, Figure S1). We constructed further synthetic contact matrices representing opening primary or secondary schools by replacing the contacts of 5-10 year-olds (primary) and 11-17 year-olds (secondary) in the ‘schools open’ contact matrix (second lockdown), with those from the ‘schools closed’ contact matrix (third lockdown) (Supplementary Figures, Figure S2).

Since the basic reproduction number scales linearly with the dominant eigenvalue of a matrix of effective contact [22], the ratio of the eigenvalues of two effective contact matrices provides a relative change in reproduction number between the three scenarios considered.

In the case where infectiousness and susceptibility are equal in all age groups, the effective contact matrix is proportional to the contact matrix itself. Under the scenarios where we assumed infectiousness and susceptibility vary with age, we converted measured contact matrices to effective contact matrices by taking the outer product of the estimated age stratified infectiousness profile and susceptibility profile vectors and calculating the eigenvalues of the Hadamard product of the resulting matrix and the contact matrices.

To demonstrate the potential impact of reopening schools, we estimated the relative increase (*k*) in reproduction number (***R***) by calculating the ratio of dominant eigenvalues of the effective contact matrix associated with the respective reopening scenario and from the current lockdown period.

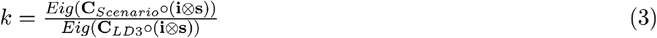

We also calculated how *R* varies from baseline values between 0.7 and 1.0, from official UK estimates of the reproduction number from [23].

## Results

### Descriptive analysis

Adults’ contacts were similar when comparing both periods of national lockdown, this is consistent across all settings and regions. Although children’s contacts at home were similar between the two periods, contacts at school and other locations were consistently higher in lockdown 2 than lockdown 3. Contacts were very similar between lockdowns in all age-group combinations other than those between children (Figure 1). For participants under 18 years-old, the mean number of contacts that were also under 18 years-old was between 6.3 (3.9 - 9.0, 90% CI) and 16.7 (13.1 - 20.4, 90% CI) across the regions of England during the November Lockdown. Such contacts were highest in South East, South West and Yorkshire and Humber and lowest in London. The mean number of contacts between children reduced to between 1.8 (1.3 - 2.5, 90% CI) and 2.6 (1.9 - 3.3, 90% CI) during the January Lockdown.

**Figure 1.**
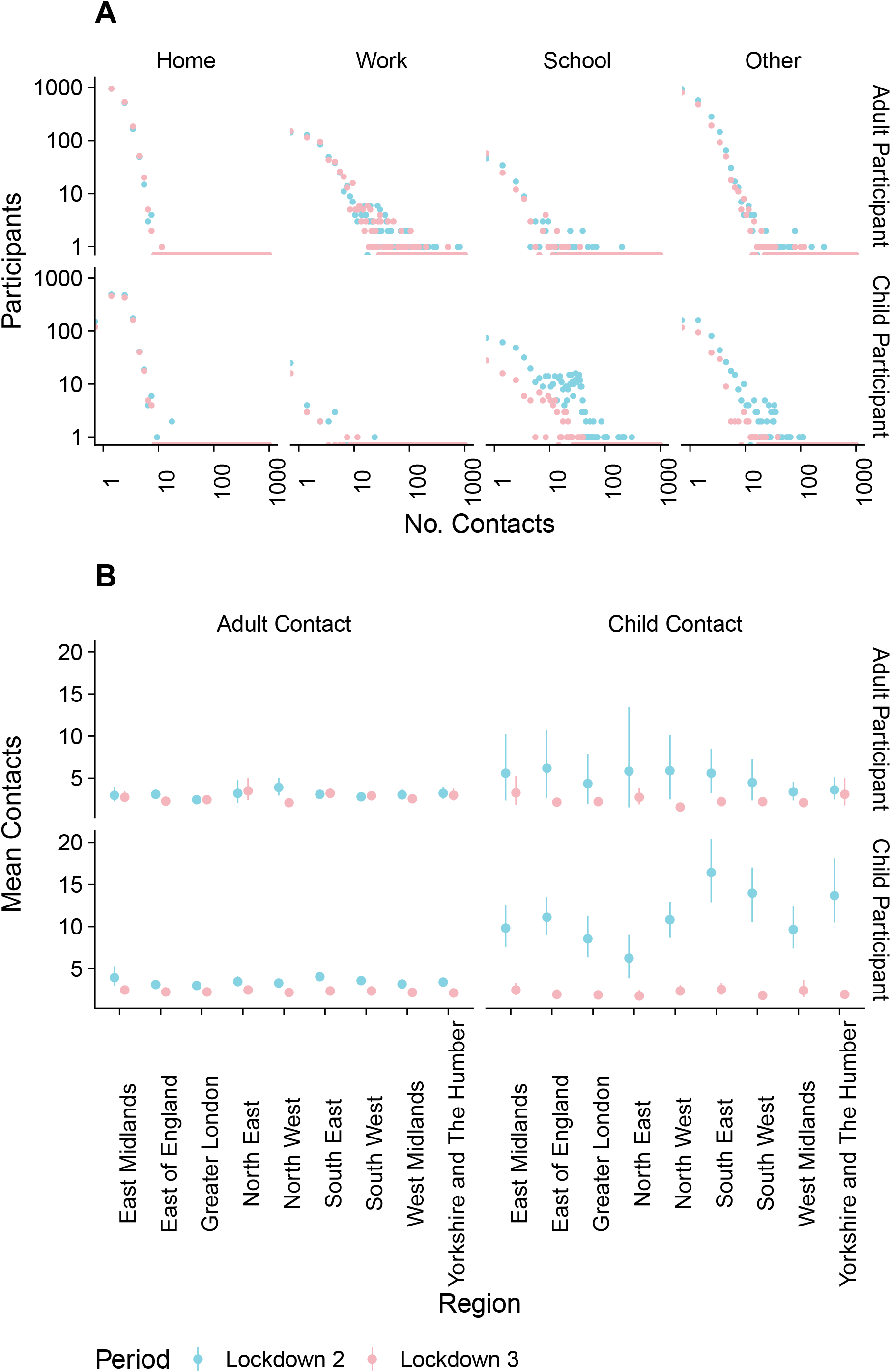
Contacts in the national lockdown periods in November (Lockdown 2) and January (Lockdown 3) . **A)** the distribution of the number of reported contacts in Home, Work, School and Other locations for Adult (> 17 years old) and Child (<= 17 years old) participants. **B)** Mean contacts reported between Children and Adults in each region of England. Error bars show the 90% CI (bootstrapped, 1000 samples).

### Estimating susceptibility in children relative to adults using CoMix data

Fitting the *R* estimates from CoMix data to time-varying *R* estimates over a period from 27th July to 10th October we estimated susceptibility of 44% (43.5% - 0.45.4%, 95% CI) in children relative to adults (Figure 2, A & C), consistent with profiles ii and iii. When we fitted from the 10th June to 10th October, 2020, we estimated 31% (29.8% - 31.4%, 95% CI) relative susceptibility in children compared to adults (Figure 2, B & D), near the lower range of ONS and Davies et al estimates. We chose to apply the second estimate as the fifth susceptibility profile (v) to represent this lower bound (Table 1) and present fits to other date ranges in the supplementary material (Supplementary Figures, Figure S4).

**Figure 2:**
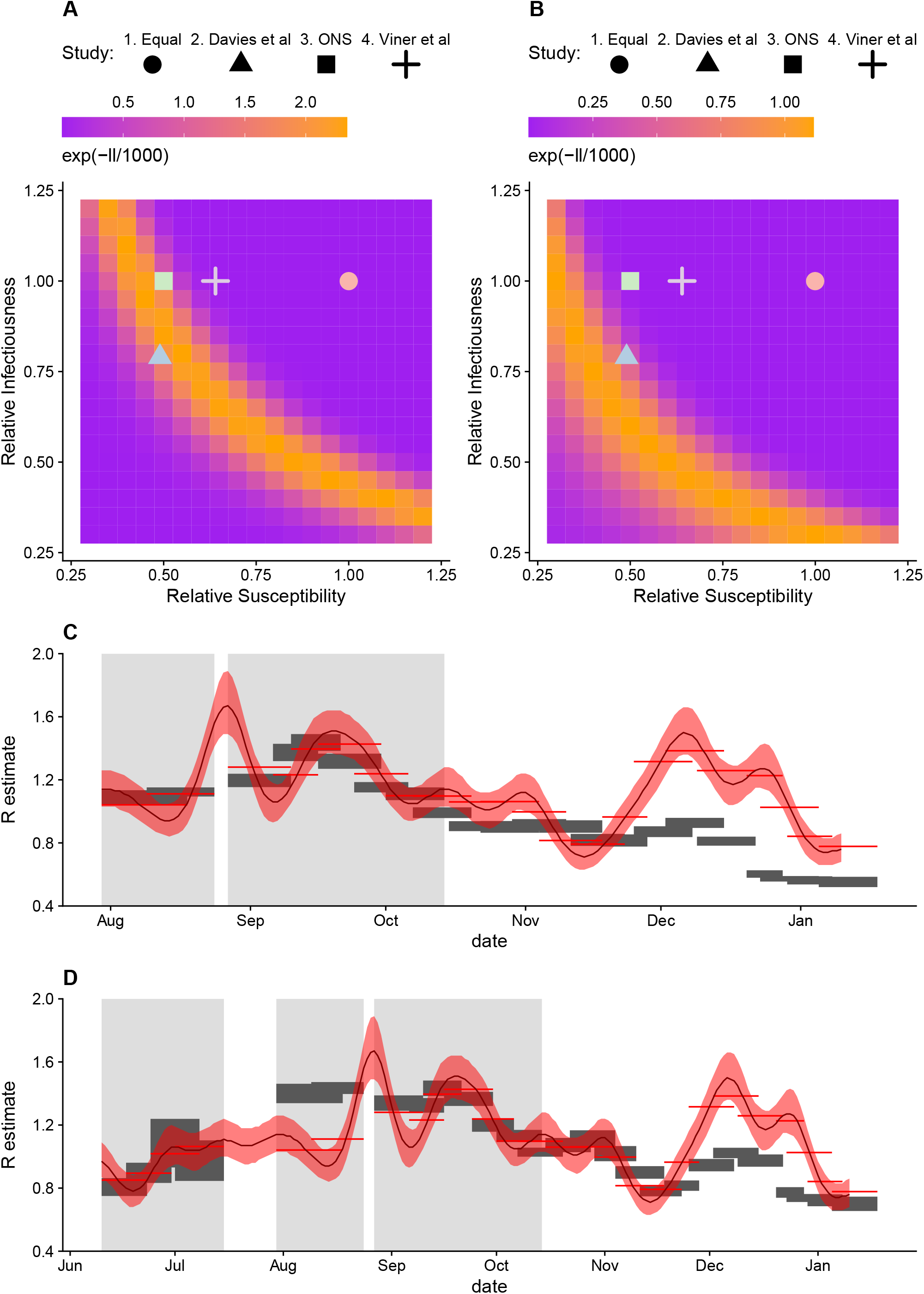
R estimates using CoMix data fit to time-varying reproduction number estimates based on the time series of cases [21]. Transformed likelihood for different combinations of relative susceptibility and infectiousness based on data from **A)** August to October and **B)** June to October and the corresponding R estimates in **C)** and **D)** respectively. 90% CI of the estimates are shown by Grey rectangles for CoMix and the red ribbon for the time-varying reproduction number estimates from case data, red bars show their mean for the CoMix survey periods. Grey shaded areas indicate fitted periods.

### Evaluation of the impact of reopening schools

Incorporating estimates of differential susceptibility and infectiousness of children compared with adults (profiles ii - v), full school reopening increased *R* by a factor of between 1.3 and 1.9 times the baseline value across the four profiles used (including 90% CI range) (Figure 3, Table 2). This would result in an increase of *R* from 0.8 to above 1.0 for these four profiles. Partial school reopening resulted in smaller increases in *R* from 0.8 to between 0.9 and 1.2.

**Figure 3:**
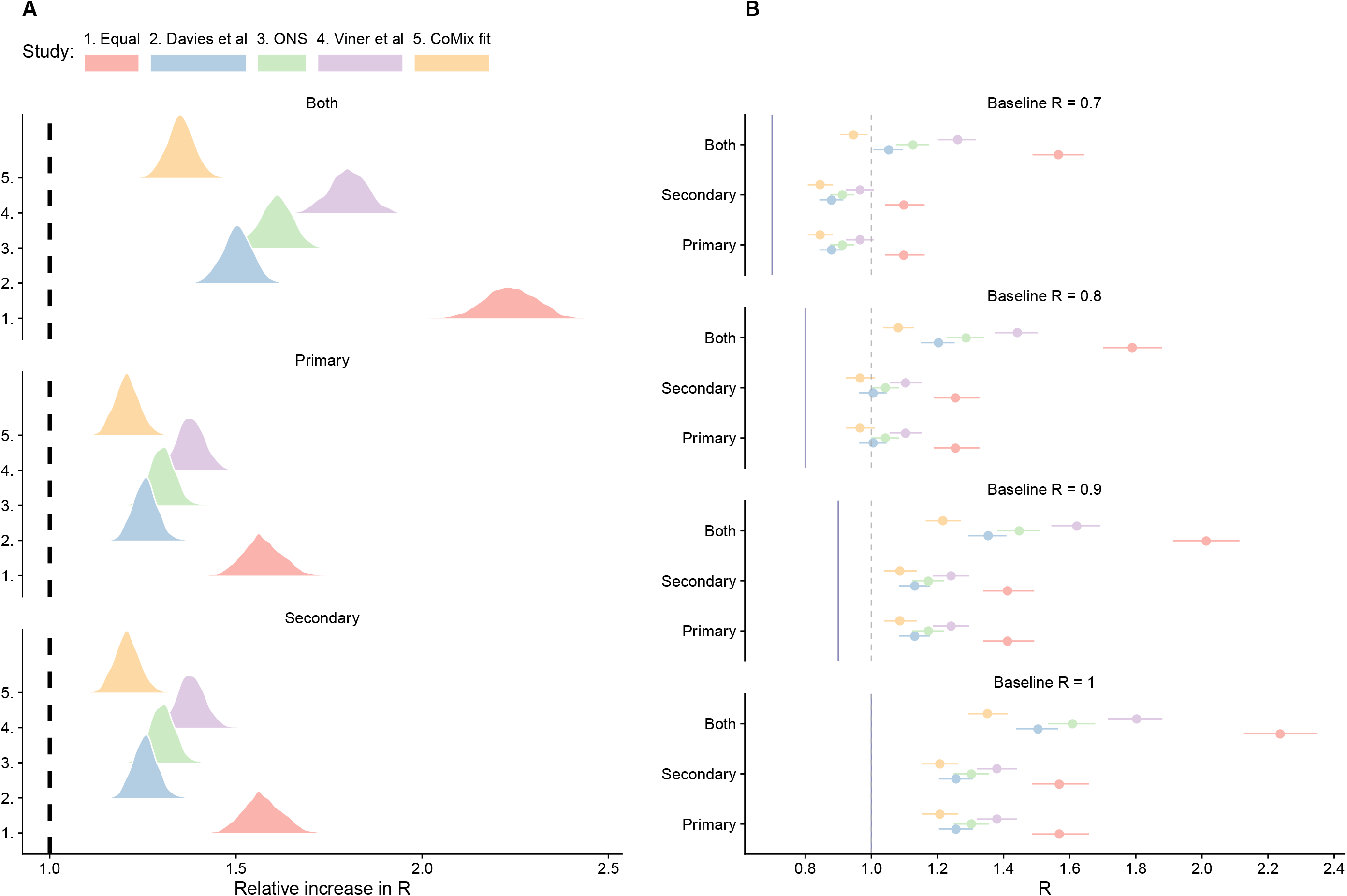
The impact of reopening schools on the reproduction number. **A)** the relative increase in R (the ratio of dominant eigenvalues between contact matrices for each reopening scenario and that for current contact patterns) under different estimates of the age profile of susceptibility and infectiousness. **B)** The estimated R after reopening schools (points, 90% CI bars) from baseline R of 0.7, 0.8, 0.9 and 1.0 (vertical line). Dashed vertical lines show R = 1.0.

**Table 2.** Expected resultant *R* if schools were reopened for different baseline values of *R* reported as median (90% CI)

| Susceptibility/<br>Infectiousness | Attendance | Baseline $R$ | | | |
| --- | --- | --- | --- | --- | --- |
|  |  | 0.7 | 0.8 | 0.9 | 1.0<br>(Scale factor) |
| **1. Equal** | Both | 1.6 (1.5 - 1.6) | 1.8 (1.7 - 1.9) | 2.0 (1.9 - 2.1) | 2.2 (2.1 - 2.3) |
|  | Primary | 1.1 (1.0 - 1.1) | 1.2 (1.2 - 1.3) | 1.4 (1.3 - 1.5) | 1.5 (1.4 - 1.6) |
|  | Secondary | 1.1 (1.0 - 1.2) | 1.3 (1.2 - 1.3) | 1.4 (1.3 - 1.5) | 1.6 (1.5 - 1.7) |
| **2. Davies et al** | Both | 1.1 (1.0 - 1.1) | 1.2 (1.1 - 1.3) | 1.4 (1.3 - 1.4) | 1.5 (1.4 - 1.6) |
|  | Primary | 0.9 (0.8 - 0.9) | 1.0 (0.9 - 1.0) | 1.1 (1.1 - 1.2) | 1.2 (1.2 - 1.3) |
|  | Secondary | 0.9 (0.8 - 0.9) | 1.0 (1.0 - 1.1) | 1.1 (1.1 - 1.2) | 1.3 (1.2 - 1.3) |
| **3. ONS** | Both | 1.1 (1.1 - 1.2) | 1.3 (1.2 - 1.3) | 1.4 (1.4 - 1.5) | 1.6 (1.5 - 1.7) |
|  | Primary | 0.9 (0.8 - 0.9) | 1.0 (1.0 - 1.1) | 1.1 (1.1 - 1.2) | 1.3 (1.2 - 1.3) |
|  | Secondary | 0.9 (0.9 - 1.0) | 1.0 (1.0 - 1.1) | 1.2 (1.1 - 1.2) | 1.3 (1.3 - 1.4) |
| **4. Viner et al** | Both | 1.3 (1.2 - 1.3) | 1.4 (1.4 - 1.5) | 1.6 (1.5 - 1.7) | 1.8 (1.7 - 1.9) |
|  | Primary | 0.9 (0.9 - 1.0) | 1.1 (1.0 - 1.1) | 1.2 (1.1 - 1.3) | 1.3 (1.3 - 1.4) |
|  | Secondary | 1.0 (0.9 - 1.0) | 1.1 (1.1 - 1.2) | 1.2 (1.2 - 1.3) | 1.4 (1.3 - 1.4) |
| **5. CoMix fit** | Both | 0.9 (0.9 - 1.0) | 1.1 (1.0 - 1.1) | 1.2 (1.2 - 1.3) | 1.4 (1.3 - 1.4) |
|  | Primary | 0.8 (0.8 - 0.9) | 0.9 (0.9 - 1.0) | 1.1 (1.0 - 1.1) | 1.2 (1.1 - 1.2) |
|  | Secondary | 0.8 (0.8 - 0.9) | 1.0 (0.9 - 1.0) | 1.1 (1.0 - 1.1) | 1.2 (1.2 - 1.3) |

When we assumed equal infectiousness and susceptibility between all age groups (profile i), reopening schools resulted in more substantial relative changes in *R*. Full school reopening increased *R* by a factor of between 2.1 and 2.3 (Figure 3, Table 2), resulting in an increase of *R* to roughly 1.7-1.9 from a baseline of 0.8 (Table 2). Partial re-opening increased *R* from 0.8 to 1.2-1.3 (Figure 3). We included these estimates for completeness but stress that assuming that children are equally infectious and susceptible as adults is not compatible with results from previous studies or our own estimates (Figure 2).

## Discussion

The potential impact of reopening schools on transmission of SARS-CoV-2 is uncertain. Although there have been many attempts to quantify the relative susceptibility and infectiousness of children and adults, these estimates need to be assessed alongside rates of contact to give an indication of the overall risk of transmission in any given setting. We combined social contact data from a large-scale survey in England during two periods of national lockdown, one with schools open and the other with schools closed, with estimates of relative susceptibility of children and adults. We used these data to quantify the potential impact of reopening schools on reproduction number.

Whereas adults’ contacts were generally similar between the two periods of lockdown, there was markedly higher contact between children during the November lockdown, when schools were open. We observed the change in contacts at school but also in other contacts outside of the home. Increased contact outside of school and home settings includes contacts in wrap around care, which would be expected to rise, however it could also indicate reduced overall adherence amongst children when attending schools physically.

The differences in contacts suggest that reopening all schools is highly likely to increase *R* above 1.0, from an assumed current value 0.8. Reopening primary or secondary is likely to increase *R* above 1.0. This would be expected to stop or reverse the fall in cases that has been observed since January 2021 [24]. The risk of cases increasing following the reopening of schools is highly dependent on the current value of *R*. Although cases of the current dominant variant (B.1.1.7) appeared to be increasing whilst national lockdown was still in place in November [10, 13], the latest national serology surveys suggest that immunity levels have substantially increased across the UK [24], resultant from both infections and the national COVID-19 vaccination program. These changes in overall immunity should be reflected in the current estimates of *R*, but these estimates are lagged due to delays in reporting [25].

In November, when schools were open, there was substantial variation in contacts between children by region. We have not presented regional estimates of the impact of reopening schools on *R* due to low numbers of observations between the lower-level age-group aggregation used in the construction of contact matrices, however the variation in mean contacts points to potential geographical variation in the impact of reopening schools, which may be lower in London than other parts of the country.

There are a number of important limitations to this work: Contacts in different settings likely contribute differently to transmission, but we assumed all contacts make equal contributions to transmission, as these differences are not well quantified in the context of control measures. If contacts at school are lower risk than those outside of school the impact of reopening schools would be lower. The age-stratified susceptibility profile is likely to change over time as natural immunity is acquired in the population. The profiles we used each reflect a single point in time. Changes in the relative immunity in children would alter the relative impact of school contacts on overall transmission. We assume adult contacts revert to those observed when all schools were open, which is conservative, in reality, particularly for partial reopening scenarios, adult contacts may not fully return to the same levels. Furthermore, there may also be differences in adherence to restrictions between the two lockdowns, unrelated to school closure. However, the change in adults’ contacts between the two periods was relatively small. The proportion of children in school varied over time due to exclusion-based control measures during the autumn, though the proportion attending school remained high during the November lockdown (Supplementary Figures, Figure S3). Contacts of children are reported by parents, which may impact their reliability, particularly in school, where parents are unlikely to witness students’ behaviour, it is unclear whether this would lead to systematic bias in reporting either more or fewer contacts.

Our work evaluates the impact of reopening schools on the reproduction number in England, which gives an indication of how transmission may be affected. However, there are other factors that reopening schools may introduce, such as the potential for children’s contact at school to provide routes of transmission between households, facilitating long chains of transmission that would be otherwise impossible[26]. We are not able to capture these network effects in this analysis, however they may play an important role in the change in epidemiology between school closure and reopening. Second, there is evidence for lower prevalence in primary school than secondary schools [8]. Our framework has not captured these differences suggesting there may be additional factors that reduce the impact of reopening primary schools relative to secondary schools. Furthermore, additional management strategies such as mass testing of school children, may serve to reduce the risk that a contact in a school results in infection beyond those implemented last year. Importantly, with the recent emergence of new variants, particularly B.1.1.7 [27], the baseline *R* will depend on proportions of these variants as well as contact patterns. Furthermore, these proportions are likely to change, potentially altering the implications of reopening schools.

Our results suggest reopening schools is likely to increase *R* close to or above 1.0, which would stop the decrease in cases observed in recent months. However, precise estimates rely heavily on the baseline values of *R* and the profiles of susceptibility, generally assuming lower susceptibility and no greater infectiousness in children relative to adults.

## Data Availability

Although it is not possible to share the contact survey data used to generate the contact matrices used in this analysis. The analysis code and contact matrices used are available in an online repository here: https://github.com/jdmunday/CoMix_schools_reopening

## List of abbreviations

CI: Confidence Interval
ONS: Office for National Statistics
UK: United Kingdom

## Declarations

### Ethics approval and consent to participate

Participation in this opt-in study was voluntary, and all analyses were carried out on anonymised data. The study and method of informed consent was approved by the ethics committee of the London School of Hygiene & Tropical Medicine Reference number 21795.

### Consent to publish

Not applicable

## Authors contributions

JDM, CIJ, WJE conceived of and planned the analysis; JDM and CIJ performed the main analysis with input from WEJ and SF; SF provided estimates of time-varying reproduction number; CIJ, KvZ, and WEJ designed the CoMix contact survey, CIJ, AG, KW, and KvZ cleaned and managed the contact survey data; All authors wrote and reviewed the manuscript. The CMMID COVID-19 Working Group provided discussion and comments.

## Acknowledgements

The authors wish to thank Dr Thomas House for his support with interpretation of the ONS susceptibility estimates. We also thank members of SPI-M for their useful discussion which helped shape the final version of this work. We would like to thank the team at Ipsos, who have been excellent in running the survey, collecting the data and allowing for the CoMix study to be implemented rapidly. Finally, we thank Katie Collis for proof reading and excellent discussions.

*The following authors were part of the Centre for Mathematical Modelling of Infectious Disease COVID-19 Working Group. Each contributed in processing, cleaning and interpretation of data, interpreted findings, contributed to the manuscript, and approved the work for publication: Yang Liu, Joel Hellewell, Nicholas G. Davies, C Julian Villabona-Arenas, Rosalind M Eggo, Akira Endo, Nikos I Bosse, Hamish P Gibbs, Carl A B Pearson, Fiona Yueqian Sun, Mark Jit, Kathleen O’Reilly, Yalda Jafari, Katherine E. Atkins, Naomi R Waterlow, Alicia Rosello, Yung-Wai Desmond Chan, Anna M Foss, Billy J Quilty, Timothy W Russell, Stefan Flasche, Simon R Procter, William Waites, Rosanna C Barnard, Adam J Kucharski, Thibaut Jombart, Graham Medley, Rachel Lowe, Fabienne Krauer, Damien C Tully, Kiesha Prem, Jiayao Lei, Oliver Brady, Frank G Sandmann, Sophie R Meakin, Kaja Abbas, Gwenan M Knight, Matthew Quaife, Mihaly Koltai, Sam Abbott, Samuel Clifford*.

## Additional Files

Supplementary Figures

