## Supplementary Figures for "Estimating the impact of reopening schools on the reproduction number of SARS-CoV-2 in England, using weekly contact survey data"

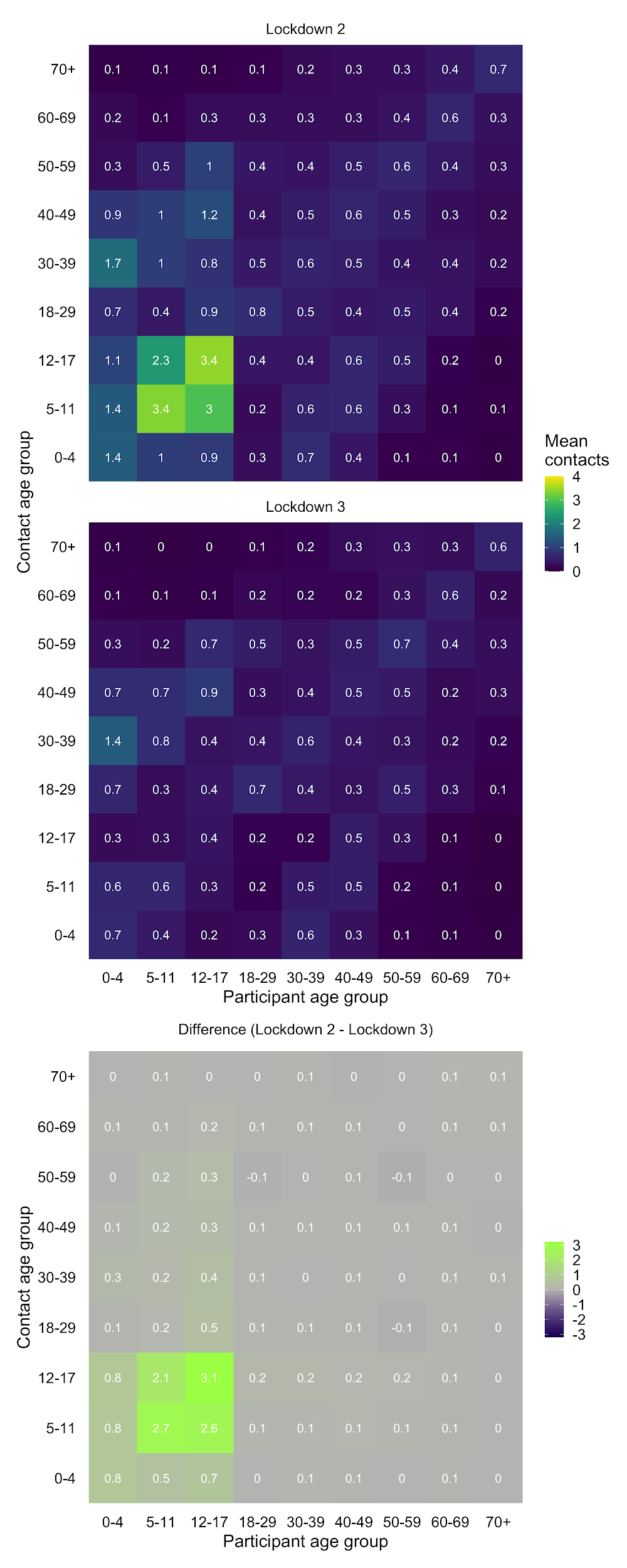

***Figure S1: Contact matrix for all contacts in England by age comparing Lockdown 2 and Lockdown 3 and the absolute difference of the cells of the matrices.*** Contacts truncated to 50 contacts per participant. Lockdown 2 data from *5th November to 2nd December 2020 and Lockdown 3 data from 5th to 18th of January 2021*

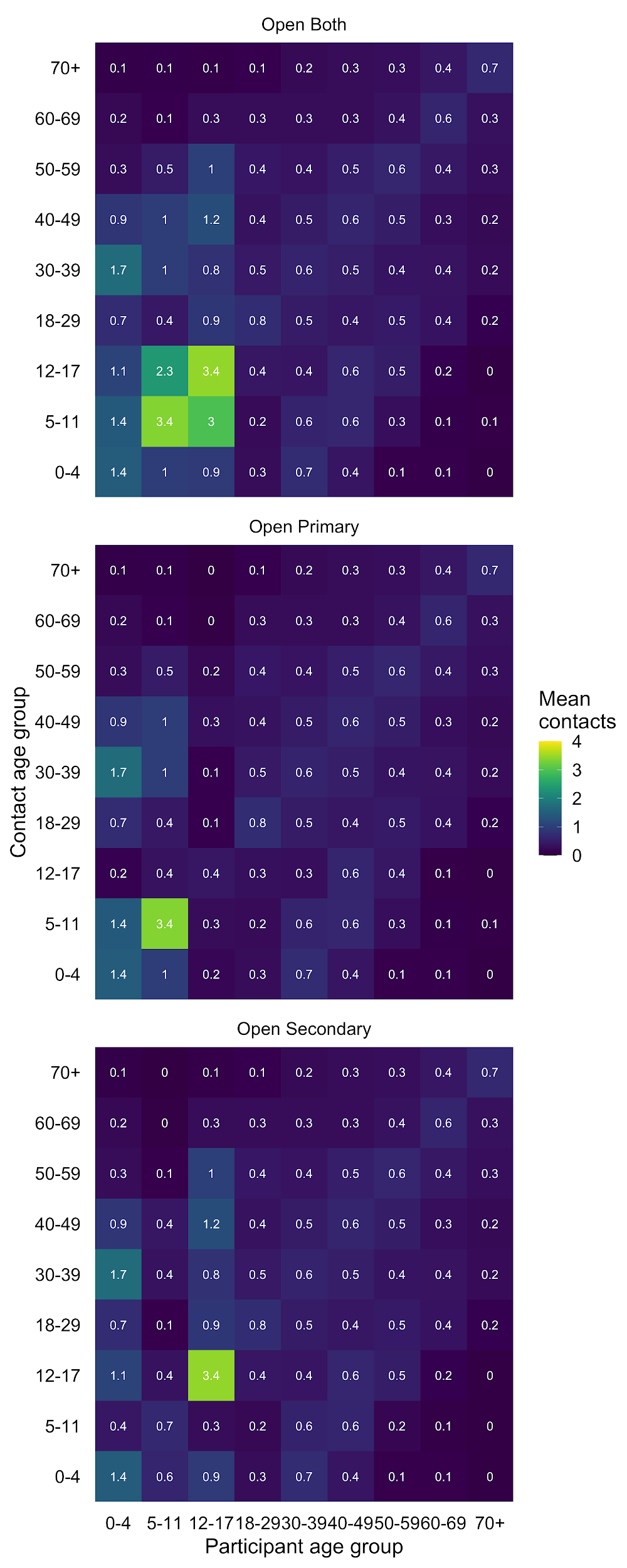

***Figure S2: Contact matrix for Scenarios included in analysis of school reopening. For all schools open the matrix calculated for Lockdown 2 was used.*** *Scenarios with Primary or Secondary schools closed replaced the 5-11 or 12-17 (respectively) column and row replaced with those calculated for Lockdown 3.* Contacts truncated to 50 contacts per participant. Lockdown 2 data from *5th November to 2nd December 2020 and Lockdown 3 data from 5th to 18th of January 2021*

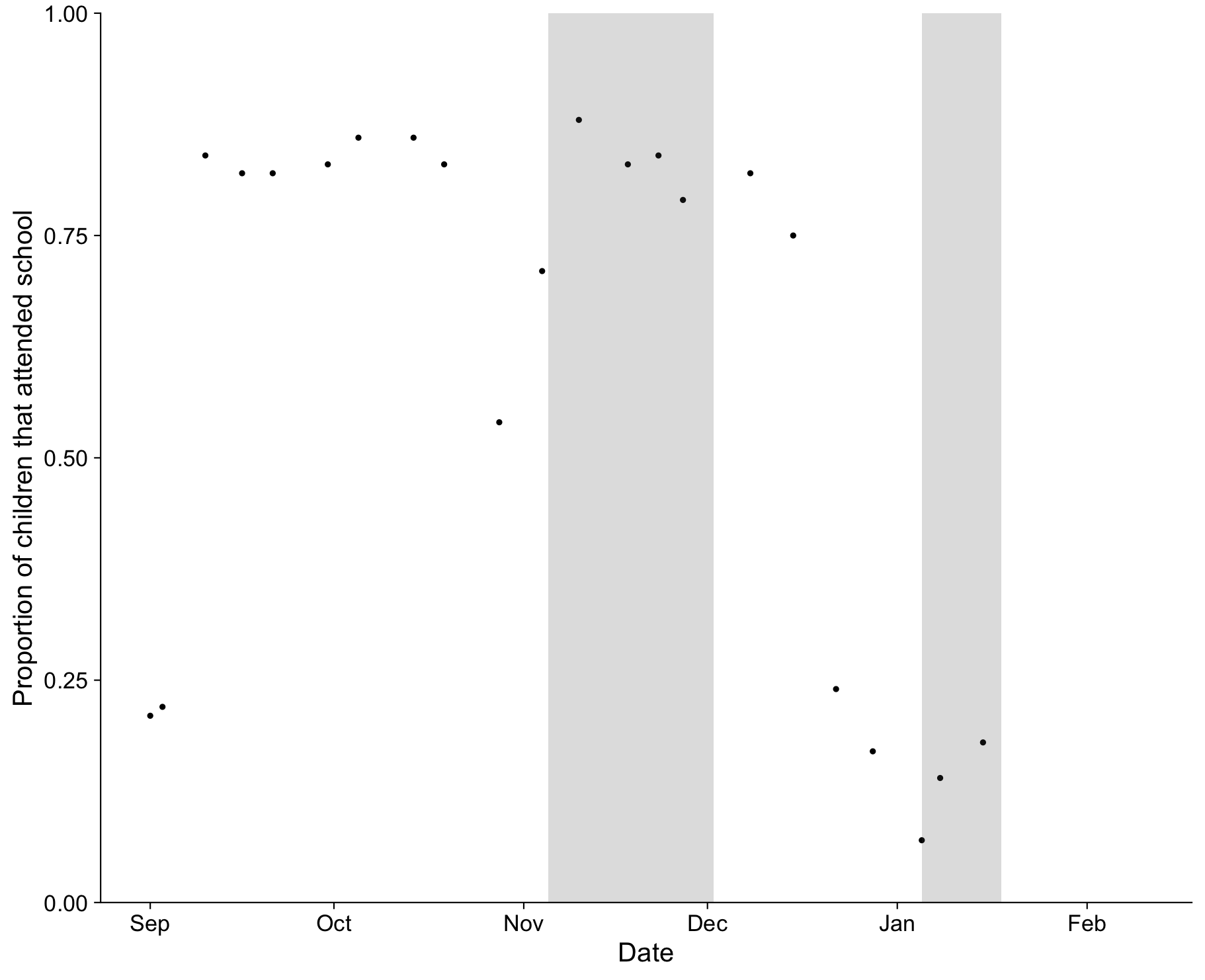

***Figure S3: The proportion of child participants who attended school on the day when contacts are recorded (with weekends removed).*** Grey bands represent the periods over which data used for this analysis was recorded (second and third lockdown).

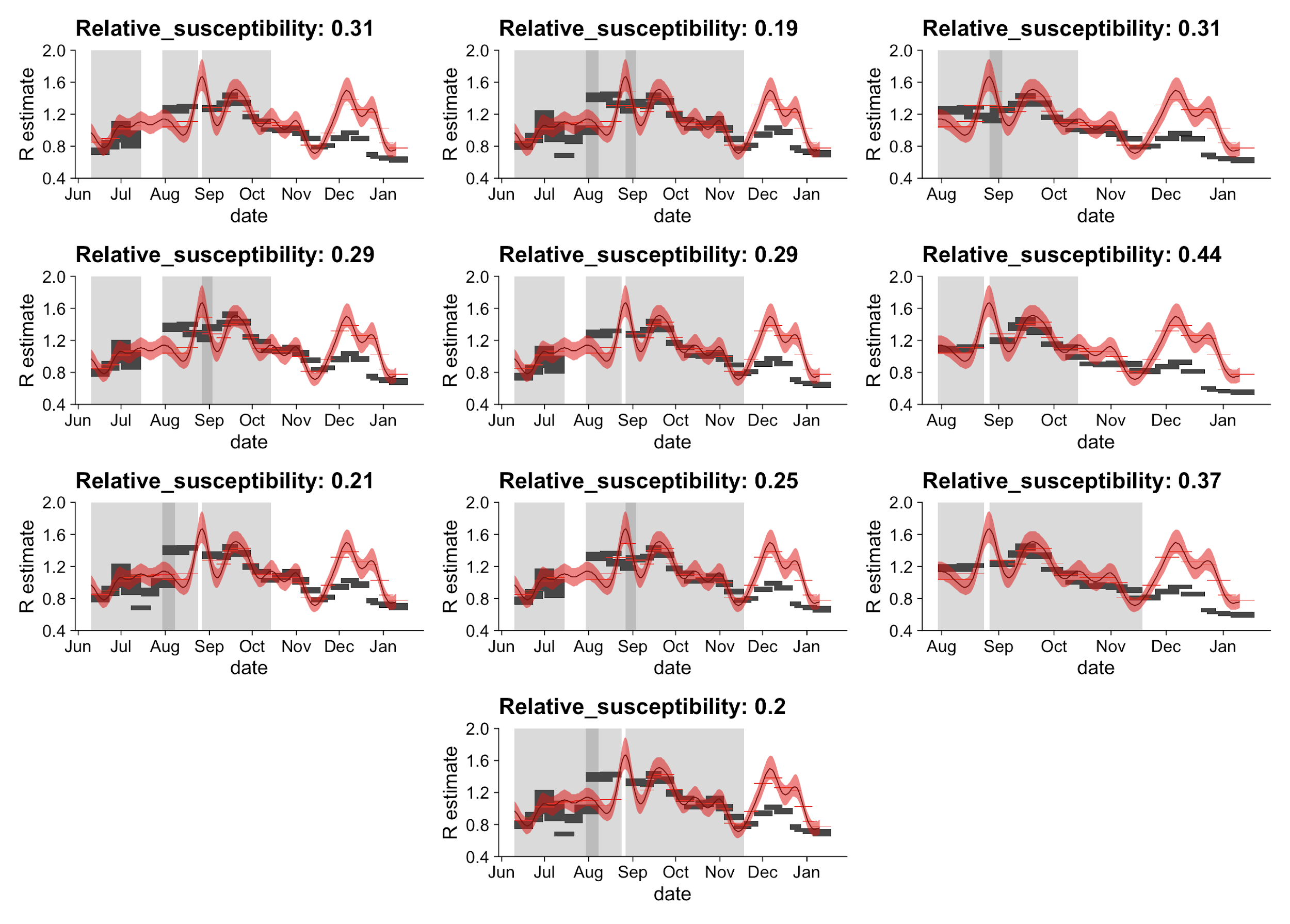

***Figure S4: Relative susceptibility found by fitting to various parts of the time-varying reproduction number estimate time series.*** *90% Confidence intervals of the estimates are shown by Grey rectangles for CoMix and the red ribbon for the time-varying reproduction number estimates from case data, red bars show their mean for the CoMix survey periods. Grey shaded areas indicate fitted periods*
